# Dermal macrophages acquire a TREM2-anchored profibrotic state in antiphospholipid syndrome that is reflected in the circulation

**DOI:** 10.64898/2026.10.29.26366773

**Authors:** Yiran Shen, Wenying Liang, Qinmengge Li, Jacqueline A. Madison, Ran Jing, Nidhi Kaza, Emily Chong, Rachael Bogle, Srilakshmi Yalavarthi, Cyrus Sarosh, Ajay Tambralli, Yu Zuo, Johann E. Gudjonsson, Hui Shi, Pei-Suen Tsou, Lam C. Tsoi, Jason S. Knight

## Abstract

Antiphospholipid syndrome (APS) is a systemic autoimmune disease best known for thrombosis and obstetric morbidity. APS is sometimes also characterized by chronic microvascular disease, which can impair renal and other organ function. Skin offers an accessible window into the tissue mechanisms underlying such manifestations. Our previous single-cell analysis of skin biopsies from three patients with established microvascular disease focused on dermal endothelial cells and implicated a Hippo–YAP1–CCN2 axis in vascular pathology. Here, we expanded single-cell profiling to dermal tissue from 30 patients with primary APS and 14 age- and sex-matched healthy controls to characterize different tissue-resident cell states and their intercellular communication networks. Cellular composition was largely preserved. Disease-associated differences were centered on myeloid transcriptional states and predicted interactions with stromal cells. Dermal myeloid cells acquired a TREM2-anchored, lipid-associated program characterized by increased expression of genes involved in lipid handling and lysosomal function. These TREM2 macrophages were the major predicted source of profibrotic osteopontin and target of stromal apolipoprotein E signaling. Expression of this dermal program correlated positively with plasma soluble TREM2 and osteopontin, and all three measures correlated inversely with estimated glomerular filtration rate. Furthermore, soluble TREM2 was higher in APS patients with suspected or established nephropathy than in those without nephropathy. Together, these findings identify a tissue macrophage program in primary APS that is reflected in the circulation and associated with renal involvement, suggesting a role for macrophage-stromal interactions in chronic microvascular disease.

## INTRODUCTION

Antiphospholipid syndrome (**APS**) is a systemic autoimmune disease in which autoantibodies targeting plasma phospholipid-binding proteins (**aPL**) produce a chronic thromboinflammatory state (1,2). Anticoagulant therapy reduces the risk of recurrent large-vessel thrombosis; however, it does not address the upstream immune dysregulation and downstream small-vessel dysfunction inherent to microvascular disease manifestations such as livedoid vasculopathy, APS nephropathy, and myocardial disease with non-obstructive coronary arteries, for which no consensus treatments exist (3,4).

The dysfunctional dermal microvasculature of APS, which presents on a spectrum from livedo racemosa to frank livedoid ulcerations, may provide a window into vascular tissue remodeling across organs (5). We previously applied single-cell RNA sequencing to the skin of three patients with APS, all with established microvascular disease and active livedo racemosa, and compared them with matched healthy volunteers. In these patients with severe disease, we identified an aberrant endothelial program centered on Yes-associated protein 1 (YAP1)-mediated signaling and connective tissue growth factor (CTGF, also known as CCN2) expression (6). This study established that APS skin undergoes biologically relevant vascular remodeling but framed it primarily around dysfunctional endothelial cells in a small cohort selected for active cutaneous disease. It is well established that systemic autoimmunity can leave disease-relevant transcriptional signatures even in relatively normal skin (7), suggesting that profiling a larger cohort of patients, whether they have active skin disease or not, could be informative.

Myeloid cells are well established to be responsive to aPL. For example, monocytes from patients with APS adopt proinflammatory and prothrombotic states (8–10). Most studies, however, have examined circulating monocytes without addressing the characteristics of monocytes and other myeloid cells within tissues.

Tissue macrophages respond to their local environment by adopting distinct transcriptional signatures. One such state is defined by high expression of triggering receptor expressed on myeloid cells 2 (**TREM2**), alongside coordinated upregulation of lipid-handling and lysosomal genes. Originally described as lipid-associated macrophages (**LAMs**) in adipose tissue (11), a similar TREM2-high state has since been identified in steatotic liver (12), atherosclerotic plaque (13), and actively remodeling atrial myocardium (14,15), where the cells may be attempting to assist in the healing process. At the same time, this state shares TREM2- and CD9-positive anchoring with the profibrotic scar-associated macrophages (SAM) inherent to liver cirrhosis (16). Of particular relevance to APS, a recent human-mouse atlas identified intrarenal monocyte and macrophage states carrying this TREM2-high program in lupus nephritis, with the identified cells also expressing apolipoprotein E (**APOE**) alongside profibrotic secreted phosphoprotein 1 (**SPP1**/osteopontin) (17). These findings link this macrophage state to autoimmune kidney disease, but its relevance to chronic microvascular remodeling in primary APS remains unknown (5,6,18,19). Although the origin of these macrophages may vary across diseases, persistent exposure to extracellular lipids and lipid-rich cellular components is a well-defined trigger, with TREM2 itself serving as the receptor for lipid-associated stimuli (11,20).

Here, we generated a single-cell transcriptomic atlas of approximately 157,000 dermal cells from 30 patients with primary APS and 14 age- and sex-matched healthy controls. Plasma and clinical data were collected at the same visit. The most striking disease-associated changes centered on predicted intercellular signaling of the myeloid compartment. Dermal myeloid cells broadly acquired a TREM2-high, LAM-like transcriptional signature. Furthermore, TREM2 macrophages were predicted to be the major dermal source of profibrotic osteopontin, which showed potential to engage and modify the function of various stromal cells. TREM2 macrophages were also the major cell type predicted to respond to APOE, which several fibroblast subpopulations prominently expressed. Finally, soluble TREM2 (**sTREM2**) and osteopontin mirrored the dermal signature as an accessible readout of this state that inversely correlated with renal function and potentially APS nephropathy across the cohort.

## METHODS

### Study participants and sample collection

We recruited 30 patients with primary APS and 14 age- and sex-matched healthy controls at the University of Michigan. All patients met the updated Sapporo classification criteria for APS. Healthy volunteers had no history of systemic autoimmune disease, current pregnancy, or active infection. At the same visit, each participant provided a 6-mm punch biopsy from the upper thigh and a blood sample; plasma was prepared from citrated blood. Nine of the 30 patients had suspected or established APS nephropathy according to the 2023 ACR/EULAR classification criteria. We extracted clinical data from the University of Michigan electronic medical record. **Table 1** provides participant characteristics. The study was approved by the University of Michigan Institutional Review Board (skin biopsy, HUM00157062; APS plasma, HUM00122519; control plasma, HUM00044257). All participants provided written informed consent before entering the study, and the study was conducted in accordance with the Declaration of Helsinki.

**Table 1.** Participants information, shown as n (%)

|  | Primary APS | Healthy controls |
|---|---|---|
| Demographics |  |  |
| Participants, n | 30 | 14 |
| Age, years, mean ± SD | 43 ± 12 | 45 ± 11 |
| Female sex, n (%) | 17 (57%) | 8 (57%) |
| White/Caucasian, n (%) | 30 (100%) | 14 (100%) |
| Hispanic or Latino, n (%) | 3 (10%) | 1 (7%) |
| Laboratory studies |  |  |
| IgG anti-β2GPI | 30 (100%) |  |
| IgM anti-β2GPI | 9 (30%) |  |
| IgG anticardiolipin | 30 (100%) |  |
| IgM anticardiolipin | 8 (27%) |  |
| Lupus anticoagulant | 28 (93%) |  |
| Triple-positive | 28 (93%) |  |
| Clinical history |  |  |
| Venous thrombosis | 16 (53%) |  |
| Arterial thrombosis | 16 (53%) |  |
| Small-vessle events | 6 (20%) |  |
| Pregnancy morbidity | 6 (35%) |  |
| Thrombocytopenia | 9 (30%) |  |
| APS nephropathy | 9 (30%) |  |
| Livedo | 14 (47%) |  |
| Skin ulcer | 6 (20%) |  |
| Obesity | 21 (70%) |  |

### Single-cell RNA sequencing library preparation, sequencing, and alignment

We incubated skin biopsies overnight in 0.4% Dispase (Life Technologies) in Hank’s balanced salt solution (Gibco) at 4°C and separated the epidermis and dermis. We kept the dermis for further library preparation, which was minced, digested in 0.2% collagenase II (Life Technologies) and 0.2% collagenase V (Sigma) in plain medium for 1.5 hours at 37°C, and strained through a 70-µm mesh. Libraries were constructed by the University of Michigan Advanced Genomics Core on the 10x Chromium platform with v3 chemistry. We sequenced libraries on an Illumina NovaSeq 6000 to generate 150-base-pair paired-end reads. Read alignment (hg38) and gene quantification were performed with Cell Ranger, and the samples were merged through seurat pipeline adjusting for ambient RNA, doublet, and batch effect..

### Single-cell quality control, clustering and subclustering

The merged expression matrix was analyzed with Seurat v4.4.0. Cells with fewer than 5^th^ or more than 95^th^ percentile detected genes or with mitochondrial content above 95^th^ percentile were excluded, and doublets were removed with scDblFinder, leaving approximately 157,000 cells. The data were log-normalized, and the top 2,000 variable genes were used for principal component analysis. The principal components explaining a cumulative 99.5% of variance were corrected for batch effects with Harmony (grouping variable: age, sex, ethnicity) and used for and UMAP visualization. Each major cell type was extracted and re-clustered with the same procedure.

### Cell-type annotation

We identified marker genes for each cluster by differential expression against all remaining cells. We manually annotated major cells and subsets according to canonical lineage markers from the literature.

### Cell composition

For each participant, the proportion of each cell population was calculated within the compartment shown in the corresponding figure panel. Proportions were compared between APS and healthy controls for each population.

### Differential expressions

Genes differentially expressed between APS and healthy controls were identified within each major cell type with the Seurat FindMarkers function (**Table S1**). Genes with an adjusted P value below 0.05 and an average log2 fold change above 0.25 were considered upregulated in APS and were used as input for the cell-cell communication analysis.

### Pseudobulk analysis

Raw counts of the pooled myeloid compartment were summed for each participant, and APS and healthy controls were compared with DESeq2 (v1.50.2). We considered genes with a Benjamini-Hochberg-adjusted P value below 0.05 to be differentially expressed. Genes were ranked by the DESeq2 Wald statistic, and gene set enrichment analysis was performed with fgsea (v1.36.2) against six MSigDB Hallmark gene sets (inflammatory response, TNF-α signaling via NF-κB, interferon gamma response, cholesterol homeostasis, fatty acid metabolism, and adipogenesis).

### Lipid-associated macrophage signature

An 18-gene LAM signature (LPL, LGALS1, LGALS3, ANXA1, TREM2, CD9, CTSB, CTSL, RGS1, FABP4, FABP5, SLC37A2, CLIC4, CD300C, NCEH1, GLIPR1, LIPA, and ATF3) was defined from published LAM gene sets (11,12). Signature scores were calculated for each cell with UCell v2.14.0 on the log-normalized expression matrix.

### Participant-level analysis

For each participant, we calculated the mean LAM score and mean expression of SPP1, TREM2, APOE, and CD44 within each myeloid or fibroblast population, and excluded participants with fewer than 10 cells in a population from the corresponding comparison. We analyzed the mean LAM score of the pooled myeloid compartment separately.

### Public datasets

Two public single-cell RNA sequencing datasets of skin from patients with systemic sclerosis (SSc) and healthy controls were reanalyzed: GSE138669 (10x Genomics) and GSE195452 (MARS-seq) (21,22). The LAM signature was scored with UCell as described above. For GSE138669. Cells with 200 to 6,000 detected genes and mitochondrial content below 20% were retained, log-normalized, integrated across samples with Harmony, and clustered at a resolution of 0.6. After removing low-quality cells and non-myeloid contaminants, the myeloid cluster was re-clustered at a resolution of 0.5, and the sub-cluster co-expressing TREM2 macrophage markers was annotated as TREM2 Mac. For GSE195452, we used the authors’ annotation (Mf and Mf_TREM2) of CD45+ sorted skin cells without re-clustering, and assigned disease status from the publicly available donor identifier.

### Cell-cell communication analysis

Cell-cell interactions were predicted separately in APS and healthy controls from known ligand-receptor pairs with CellPhoneDB (v5.0.0) and CellChat (v2.2.0.9001) (23,24). For CellPhoneDB, we applied the differentially expressed gene-based method to the 12 major cell types, using genes upregulated in each group as input. For CellChat, interactions among endothelial, myeloid, fibroblast, and pericyte subtypes were inferred with the human CellChatDB, and the APS and healthy control results were merged for comparison.

### Plasma ELISAs

Plasma sTREM2, sTREM1, osteopontin, and sCD163 were measured with R&D Systems kits (Human TREM2 DuoSet DY1828-05, 1:10; Human TREM-1 Quantikine DTRM10C, 1:2; Human Osteopontin Quantikine DOST00, 1:25; Human CD163 Quantikine DC1630, 1:25) according to the manufacturer’s instructions.

### Statistical analysis and software

We calculated cell proportions, signature scores, and gene expression for each participant and compared them between APS and healthy controls using two-sided Mann-Whitney tests. We used the Kruskal-Wallis test with Dunn’s multiple comparison test for three-group comparisons, and assessed associations with Spearman’s correlation in GraphPad Prism 11. Single-cell analyses were performed in R v4.5.1 with Seurat v4.4.0. Partial Spearman correlations controlling for BMI were calculated with the R package ppcor (v1.1). P values were adjusted by the Benjamini-Hochberg method within each figure panel, and q < 0.05 was considered statistically significant.

### Data availability

All study data will be made available upon reasonable request.

## RESULTS

### A single-cell atlas of APS dermis localizes disease-associated signaling to the myeloid compartment

We profiled upper-thigh skin biopsies from 30 patients with primary APS and 14 age- and sex-matched healthy controls by droplet-based single-cell RNA sequencing (**Figure 1A**, **Table 1**). Paired blood specimens and clinical data collected at the same study visit. Of the 30 patients with APS, 14 had a history of livedoid skin changes and six a history of ulcerative skin disease, though none of the biopsies were from active ulcers. After quality control, approximately 157,000 dermal cells were resolved into 12 major cell types, annotated by canonical lineage markers (**Figure 1B–C**).

**Figure 1.**
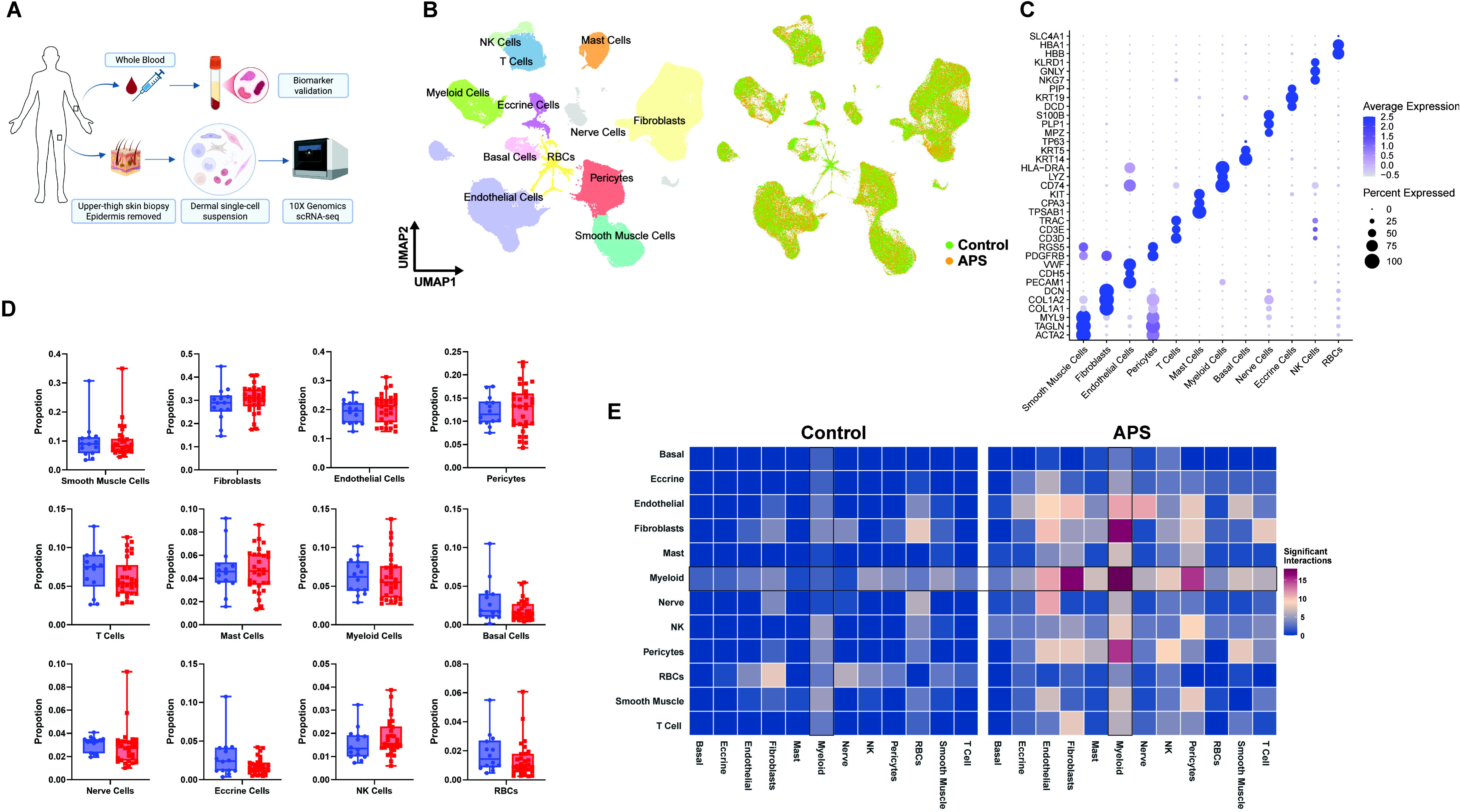
Single-cell atlas and intercellular communication landscape of APS dermis. (A) Study design and biospecimen framework. Dermal cells were isolated from upper-thigh skin and profiled by droplet-based single-cell RNA sequencing. The cohort included 30 patients with primary antiphospholipid syndrome (APS) and 14 age- and sex-matched healthy controls (HCs). Paired blood-derived specimens and clinical data were collected at the same study visit, as indicated. (B) Uniform manifold approximation and projection (UMAP) of approximately 157,000 quality-controlled dermal cells, colored by 12 major cell-type annotations. The corresponding HC and APS UMAPs are shown on the right. (C) Dot plot showing representative canonical markers used to annotate the 12 major dermal cell types. (D) Sample-level proportions of the 12 major cell types in APS and HC skin. Each point represents one participant. The horizontal line within the box represents the median, the whiskers extend from the minimum to the maximum values, and the box boundaries represent the 25th and 75th percentiles. APS and HC were compared separately for each cell type using two-sided Mann–Whitney tests, followed by Benjamini–Hochberg (BH) correction across the 12 comparisons. No major cell type differed significantly between APS and HC after correction (q > 0.05). (E) CellPhoneDB analysis of candidate intercellular communication in HC and APS dermis. Within each annotated cell population, significantly differential expression between APS and HC was assessed using Seurat FindMarkers and supplied to the CellPhoneDB workflow.

The proportions of all 12 major cell types were similar between APS and healthy controls after Benjamini–Hochberg (BH) correction (q > 0.05; **Figure 1D**). Since cell abundance alone did not distinguish APS from control dermis, we investigated the transcriptional states and predicted intercellular signaling that these cells adopt in situ. Within each annotated population, we assessed APS-versus-control differential expression (**Table S1**) and then used CellPhoneDB to enumerate significant ligand–receptor interactions in each group (**Figure 1E**). In healthy control dermis, predicted interactions were relatively sparse and broadly distributed. In contrast, the interaction landscape of APS dermis was denser and particularly enriched for the myeloid compartment. Myeloid cells formed the most connected hub and showed the largest gains in signaling with fibroblasts, endothelial cells, and pericytes (**Figure 1E**). Together, these data indicate that APS dermis remodels at the level of intercellular communication rather than cellular composition, with the myeloid compartment emerging as a principal hub of this rewiring.

We next asked how APS reshapes myeloid cell gene expression. In sample-level pseudobulk differential expression analysis of the total myeloid compartment, the genes most strongly upregulated in APS were associated with lipid handling. Gene set enrichment analysis demonstrated coordinated upregulation of both lipid–metabolic (fatty acid metabolism, cholesterol homeostasis) and inflammatory (tumor necrosis factor/NF-κB, interferon gamma) programs (**Figure S1A**). Prominent differentially expressed genes for the former included apolipoprotein C1 (*APOC1*), fatty acid-binding proteins 4 and 5 (*FABP4* and *FABP5*), lipoprotein lipase (*LPL*), lysosomal acid lipase (*LIPA*), and *TREM2*, all with BH-adjusted p < 0.05 (**Figure S1B**). Since the inflammatory programs were not reflected by individually prominent genes, the most meaningful transcriptional changes in APS myeloid cells appeared to center on lipid metabolism, prompting us to identify which myeloid subpopulations carried such gene expression changes.

### APS dermal myeloid cells acquire a LAM-like program, which is most prominent in TREM2 macrophages

We subclustered the dermal myeloid compartment into five transcriptionally distinct populations: M1-like macrophages, M2-like macrophages, TREM2 macrophages, conventional type 1 dendritic (**cDC1**) cells, and cDC2B cells, each defined by canonical markers (**Figure 2A–B**). The TREM2 macrophage population expressed the scavenger receptor CD163 but not TREM1, discriminating it from a traditional proinflammatory macrophage phenotype (**Figure S2A**) (25). As with the major cell types, proportions of these five myeloid subpopulations were comparable in APS and healthy controls (q > 0.05; **Figure 2C**).

**Figure 2.**
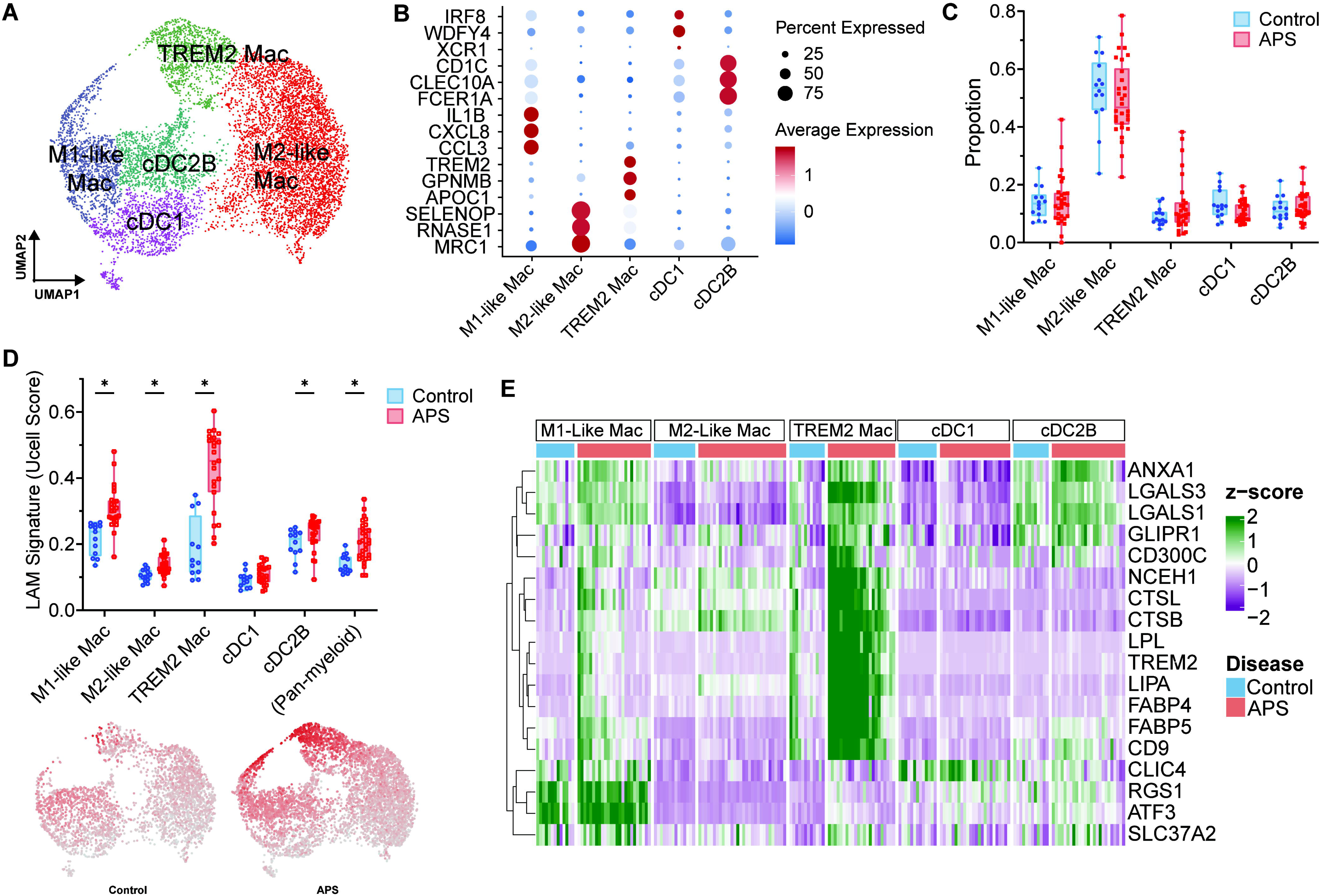
APS dermal myeloid cells acquire a TREM2-anchored lipid-associated macrophage program. (A) UMAP of the dermal myeloid compartment, resolving five transcriptionally defined populations: M1-like inflammatory macrophages, M2-like resident macrophages, TREM2 macrophages, cDC1 cells, and cDC2B cells. (B) Dot plot showing representative markers used to annotate the five myeloid populations. (C) Sample-level proportions of the five myeloid populations in APS and HC skin. Each point represents one participant. No myeloid population met the prespecified false discovery rate threshold (q > 0.05). (D) Increased lipid-associated macrophage (LAM) signature activity in APS myeloid cells. The upper panel shows sample-level mean UCell scores for a predefined 18-gene LAM signature within each myeloid population and across the complete pan-myeloid compartment. TREM2, M1-like inflammatory macrophages, M2-like resident macrophages, and cDC2B cells met the prespecified false discovery rate threshold (*), whereas cDC1 cells did not (q = 0.0829). The pan-myeloid score was analyzed separately (two-sided Mann–Whitney P=0.0003). The lower panel projects per-cell LAM UCell scores onto the shared myeloid UMAP embedding, displayed separately for HC and APS. nd: not a discovery. (E) Sample-level expression of the 18 LAM signature genes across the five myeloid populations. Columns represent individual participants grouped by myeloid population and disease status, and rows represent LAM signature genes (row-normalized).

We next scored all five myeloid populations with a prespecified 18-gene LAM signature, comparing sample-level scores between APS and healthy controls (11,12). Expression of the signature was higher in APS across the entire myeloid compartment (Pan-myeloid, p = 0.0003) and in most individual myeloid subpopulations, reaching significance in M1-like, M2-like, and TREM2 macrophages, as well as in cDC2B, but not cDC1 cells (**Figure 2D**). Although the LAM signature was elevated across multiple myeloid subpopulations, the central lipid-handling and lysosomal program was particularly strong in APS TREM2 macrophages (**Figure 2E**). In addition to the canonical LAM surface markers TREM2 and CD9, APS TREM2 macrophages upregulated genes for extracellular lipolysis (*LPL*), lipid transport (*FABP4*, *FABP5*), and lysosomal hydrolysis (*LIPA*, *NCEH1*, *CTSB*, *CTSL*). Together, these data indicate that APS dermal myeloid cells broadly acquire a LAM signature, with the TREM2 macrophage representing its prototypical and most fully expressed state.

### SPP1/osteopontin- and APOE-mediated signaling links dermal TREM2 macrophages to the stroma in APS

The intensity of predicted signaling between myeloid cells and fibroblasts (**Figure 1E**) suggested that the LAM-like signature, most prominently expressed by TREM2 macrophages (**Figure 2E**), might support communication with the stromal compartment. As the largest dermal cell populations, we subclustered fibroblasts into eight transcriptionally distinct populations: CCL19+, COL11A1+, COL15A1+, COL18A1+, PI16+, PLA2G2A+, NFKB+, and SFRP2+, each defined by canonical markers (**Figure 3A–B**) (26). We also separated the endothelial and pericyte compartments into their respective subpopulations (**Figure S3A–F**). Similar to the myeloid compartment, cell composition was not significantly different between APS and healthy controls for any fibroblast (**Figure 3C**), endothelial, or pericyte (**Figure S3C–F**) subpopulation.

**Figure 3.**
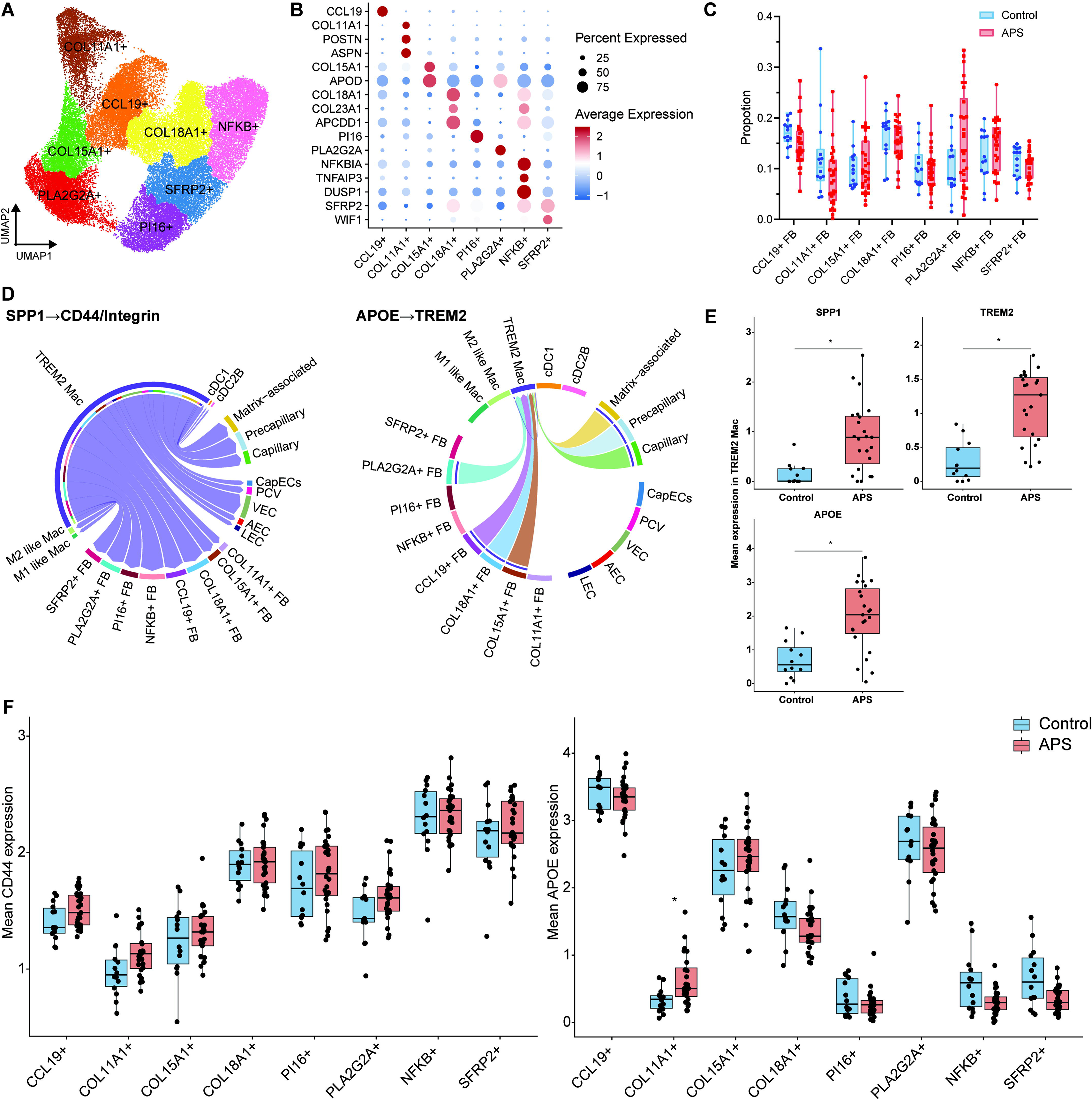
Predicted SPP1 (osteopontin) –CD44/integrin and APOE–TREM2 signaling connects TREM2 macrophages with fibroblast states in APS dermis. (A) UMAP of the integrated dermal fibroblast compartment, resolving eight transcriptionally defined populations: CCL19+, COL11A1+, COL15A1+, COL18A1+, PI16+, PLA2G2A+, NFKB+, and SFRP2+ fibroblasts. (B) Dot plot showing representative markers used to annotate the eight fibroblast populations. (C) Sample-level proportions of the eight fibroblast populations in APS and HC skin. Each point represents one participant. No fibroblast population met the prespecified false discovery rate threshold (q>0.05). (D) Focused chord diagrams of CellChat SPP1 (osteopontin) –CD44/integrin signaling (left) and APOE–TREM2 signaling (right) among myeloid, fibroblast, endothelial, and perivascular cell populations in APS dermis. Ribbons indicate predicted communication from source to target populations. SPP1 signaling was broadly directed from TREM2 macrophages to fibroblast, endothelial, pericyte, and other myeloid populations, whereas APOE signaling was directed to TREM2 macrophages from fibroblast and pericyte populations. Both signaling networks were detected in APS but not in HC. (E) Sample-level mean expression of SPP1, TREM2, and APOE within TREM2 macrophages. Participants contributing fewer than 10 TREM2 macrophages were excluded, resulting in 23 APS and 12 HC participants for each comparison. SPP1, TREM2, and APOE were significantly increased in APS TREM2 macrophages after BH correction (q < 0.05). (F) Sample-level mean expression of CD44 (left) and APOE (right) across the eight fibroblast populations. Each point represents one evaluable participant; the number of participants varied according to the availability of cells from the corresponding fibroblast population (n > 10). CD44 did not differ significantly between APS and HC in any fibroblast population after BH correction, and APOE was selectively increased in COL11A1+ fibroblasts in APS (q=0.0456).

Across myeloid, fibroblast, endothelial, and pericyte subpopulations, TREM2 macrophages showed the largest change in the number of predicted interactions between APS and healthy controls (**Figure S3G**). To identify the most relevant connections, we ranked the predicted strength of the contributing ligand–receptor pairs. SPP1 (also known as osteopontin) and APOE were the highest-ranked ligands, both carrying significant information flow in APS skin, but not in healthy control skin (**Figure S3H**). In this ligand–receptor interaction analysis, TREM2 macrophages were the major predicted source of osteopontin, which was predicted to engage CD44 on fibroblast and pericyte populations, and integrin receptors broadly across fibroblast, endothelial, pericyte, and other myeloid populations. TREM2 macrophages were also the major predicted target of APOE, which was contributed by fibroblast and pericyte subpopulations and expected to interact with the TREM2/TYRO protein tyrosine kinase-binding protein (TYROBP) receptor complex (**Figure 3D**).

Finally, we examined whether gene expression underlying these predicted interactions was altered in APS at the sample level. Compared with healthy controls, *SPP1* gene expression was significantly increased in APS TREM2 macrophages (q < 0.05; **Figure 3E**), whereas *CD44* expression did not differ in any fibroblast population (**Figure 3F**). Furthermore, *TREM2* gene expression was increased in TREM2 macrophages (**Figure 3E**), whereas *APOE* expression did not increase in the fibroblast populations predicted to contribute it as a ligand (**Figure 3F**). Interestingly, TREM2 macrophages also increased their own *APOE* expression in APS (**Figure 3E**) and were therefore among its predicted sources (**Figure 3D**).

### Circulating sTREM2 mirrors the dermal LAM signature and inversely associates with renal function in APS

To ask whether the prespecified 18-gene dermal myeloid LAM signature has a circulating correlate, we assessed plasma collected alongside the biopsy samples (**Figure 1A**). The myeloid LAM signature positively correlated with plasma sTREM2 (ρ = 0.60, P = 0.0005) and plasma osteopontin (ρ = 0.54, P = 0.0021; **Figure 4A**). We also asked whether similar correlations might be observed with other myeloid activation markers. CD163 and TREM1 are such markers, though they are not significantly expressed by TREM2 macrophages (**Figure S2A**). Neither soluble CD163 (ρ = 0.15; P = 0.43; **Figure S4A**) nor soluble TREM1 (ρ = 0.24; P = 0.20; **Figure S4B**) in plasma correlated with the myeloid LAM signature.

**Figure 4.**
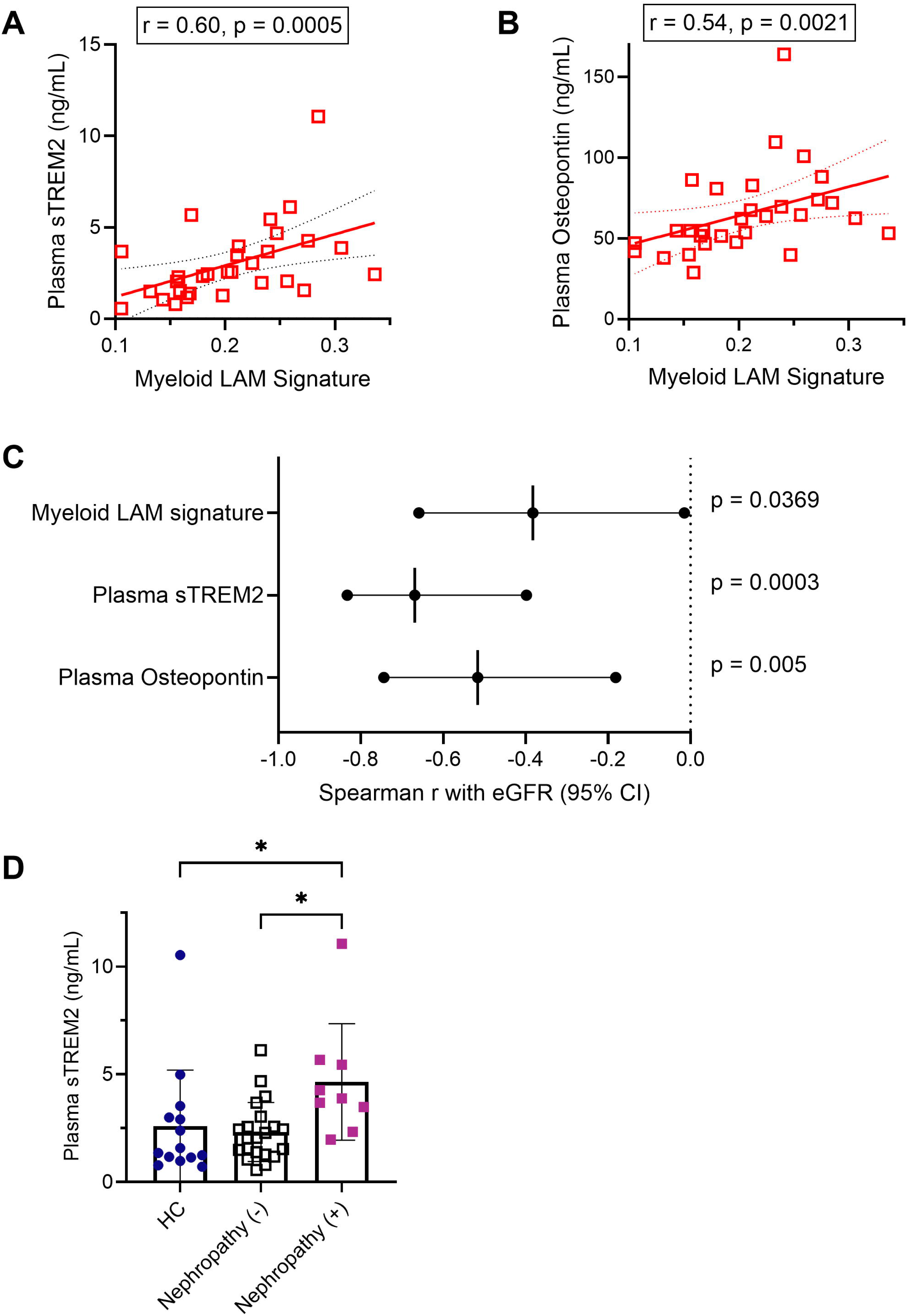
The dermal LAM program is reflected in paired plasma biomarkers and associates with renal function in APS. (A) Associations between the tissue myeloid lipid-associated macrophage (LAM) signature and paired plasma concentrations of soluble TREM2 (sTREM2; left) and osteopontin (encoded by SPP1; right) among 30 patients with APS. Each point represents one participant sampled concurrently for skin and plasma. Associations were evaluated by two-sided Spearman correlation. The LAM signature correlated positively with plasma sTREM2 (r = 0.60, 95% CI 0.2955–0.7937, P = 0.0005) and plasma OPN (r = 0.54, 95% CI 0.2113–0.7579, P = 0.0021). (B) Forest plot showing associations of LAM signature, plasma sTREM2, and plasma osteopontin with estimated glomerular filtration rate (eGFR) among 30 patients with APS. Points indicate Spearman correlation coefficients, and horizontal lines indicate 95% confidence intervals. LAM signature (r= -0.3826, 95% CI, −0.6592 to −0.01475; q=0.0369), plasma sTREM2 (r= -0.6692, 95% CI, −0.8329 to −0.3978; q<0.0003), and plasma SPP1 (r= -0.5165, 95% CI, −0.7442 to −0.1812; q=0.00525) were inversely associated with eGFR. (C) Plasma sTREM2 in patients with APS compared to the HCs, and stratified by documented nephropathy (nephropathy+, n = 9; nephropathy−, n = 21) and in HCs (n = 14). Plasma sTREM2 was elevated in nephropathy+ versus nephropathy−.

Finally, because both cutaneous and renal manifestations of APS are examples of microvascular disease (27), we asked whether the markers assessed above were associated with renal function. All three of the myeloid LAM signature, plasma sTREM2, and plasma osteopontin were inversely correlated with estimated glomerular filtration rate (**eGFR**) across the APS cohort (**Figure 4B**). We repeated these analyses as partial correlations controlling for BMI; the correlation coefficients changed by less than 0.02 and all three associations remained significant (**Table S2**). Among the 30 APS patients, nine had either suspected or established APS nephropathy according to the 2023 ACR/EULAR classification criteria. Although plasma sTREM2 did not differ between APS patients and healthy controls overall (p = 0.15), it was elevated in nephropathy-positive patients relative to both nephropathy-negative patients and controls (**Figure 4C**), positioning it as a within-disease correlate of renal involvement. Neither the tissue LAM signature nor plasma osteopontin distinguished nephropathy-positive from nephropathy-negative patients (**Figure S4C**) in this analysis.

## DISCUSSION

By profiling upper-thigh skin biopsies from patients with primary APS and matched healthy controls, we found that dermal alterations in APS were especially prominent in the myeloid compartment. In particular, we identified a TREM2-anchored LAM signature across APS myeloid cells, somewhat reminiscent of tissue adaptation in other chronic disease contexts (11–16), including in a recent study of lupus nephritis (17). Furthermore, this tissue macrophage state had plasma correlates that tracked inversely with renal function, raising the possibility that these myeloid changes are not confined to the skin. Together, these observations indicate that tissue macrophages in APS acquire a LAM-like, stromal-interactive, likely profibrotic state with potential to amplify chronic APS microvascular manifestations.

The LAM phenotype is a conserved cell state defined across tissues by lipid-handling and lysosomal genes, including *TREM2*, *APOE*, and, when taking on profibrotic characteristics, *SPP1* (11,28). Interestingly, this acquisition was not restricted to TREM2 macrophages in our analysis, suggesting that the signals inducing this state act across myeloid populations in the dermal microenvironment, although the central lipid-handling and lysosomal program was most fully expressed in TREM2 macrophages. Cross-sectional profiling cannot resolve whether this state arises from adaptation of resident dermal macrophages or from in situ differentiation of recruited monocytes.

Dermal cell–cell communication modeling identified TREM2 macrophages as the major predicted source of osteopontin and the major predicted target of APOE in stromal signaling. Through osteopontin, TREM2 macrophages were predicted to engage CD44 and integrins across fibroblast, endothelial, and pericyte populations, and *SPP1* gene expression was increased in APS TREM2 macrophages, consistent with its established roles in stromal cell migration, myofibroblast activation, and tissue fibrosis (29–36), all features of chronic APS nephropathy (18,37). Conversely, the APOE axis directed signals from fibroblast and pericyte populations toward APS TREM2 macrophages, where *TREM2* gene expression was increased. In both axes, disease-associated changes localized to TREM2 macrophages, whereas CD44 and APOE gene expression in their predicted stromal partners was unchanged. Interestingly, TREM2 macrophages were among the predicted sources of APOE, while also demonstrating increased *APOE* gene expression in APS, suggesting the potential for a significant autocrine contribution to this axis.

Plasma sTREM2 and osteopontin correlated with the tissue LAM signature, whereas other macrophage activation markers, including soluble CD163 and soluble TREM1, did not. These circulating proteins therefore appear to reflect a particular macrophage state rather than generalized myeloid activation. Because microvascular narrowing and occlusion underlie both cutaneous and renal manifestations of APS (18,34), we assessed eGFR and found that the tissue LAM signature, plasma sTREM2, and plasma osteopontin were each inversely correlated with it. Across the three measures, only plasma sTREM2 separated the nine patients with nephropathy from those without. TREM2 is shed primarily from macrophages (38), whereas plasma osteopontin can come from neutrophils and other sources, which may obscure its link to a macrophage state in a given organ. Furthermore, with only nine patients, this comparison remains exploratory.

Amongst rheumatic diseases, systemic sclerosis may provide the most relevant comparator for chronic dermal microvascular disease. We therefore applied the same 18-gene LAM signature to two published systemic sclerosis skin datasets (GSE138669, GSE195452) (21,22). As expected, we found evidence of a similar TREM2-anchored, LAM-like signature in dermal macrophage subsets of both datasets, although not significantly different from healthy controls, indicating that this state is not wholly unique to APS (**Figure S5A–D**). However, because cell recovery per donor was not comparable across these datasets (**Figure S5E**), only our biopsies yielded enough TREM2 macrophages to support donor-level analysis, allowing us to link the tissue observation to the clinical variables described above.

The broad expression of the LAM-like signature across myeloid subpopulations, without marked numerical expansion, suggests sustained receptor activation within the dermal perivascular microenvironment. Molecularly, TREM2 functions as a cell-surface myeloid receptor that can engage distinct classes of disease-relevant danger signals. Prominent examples include anionic and oxidized phospholipids exposed on damaged membranes, as well as myeloperoxidase presented on neutrophil extracellular traps (**NETs**) (20,39–41). In APS, persistent microvascular injury exposes anionic phospholipid surfaces likely to capture pathogenic APS-relevant autoantigens, including beta-2 glycoprotein I (3,42), while ongoing NETosis may provide a continuous extracellular source of myeloperoxidase (43). Together, the chronic availability of membrane-associated lipids alongside neutrophil-derived enzymes are candidate endogenous inputs that might sustain this localized macrophage activation state.

An important limitation of our study is its cross-sectional sampling. The relationship between this macrophage state and progressive microvascular disease will only be able to be resolved through longitudinal assessments. Understanding the signals that sustain this signaling will also likely require developing new experimental models that go beyond acute thrombosis. Going forward, large prospective cohorts should seek to establish the value of plasma sTREM2 and osteopontin for tracking, and potentially diagnosing, microvascular involvement. At the same time, direct profiling of organs beyond the skin, most notably the kidneys, should be a priority. The hope is that further defining this TREM2-anchored stromal biology will offer a route toward mechanism-directed intervention in the non-thrombotic manifestations of APS, for which treatment options remain limited.

## ACKNOWLEDGMENTS

Portions of this work were supported by the assistant secretary of defense for health affairs, endorsed by the Department of Defense through the Lupus Research Program under award number W81XWH-22-1-0895 (to Dr Knight). The opinions, interpretations, conclusions, and recommendations contained herein are those of the author(s) and are not necessarily endorsed by the Department of Defense.

