## Supplemental Figures for "Dermal macrophages acquire a TREM2-anchored profibrotic state in antiphospholipid syndrome that is reflected in the circulation"

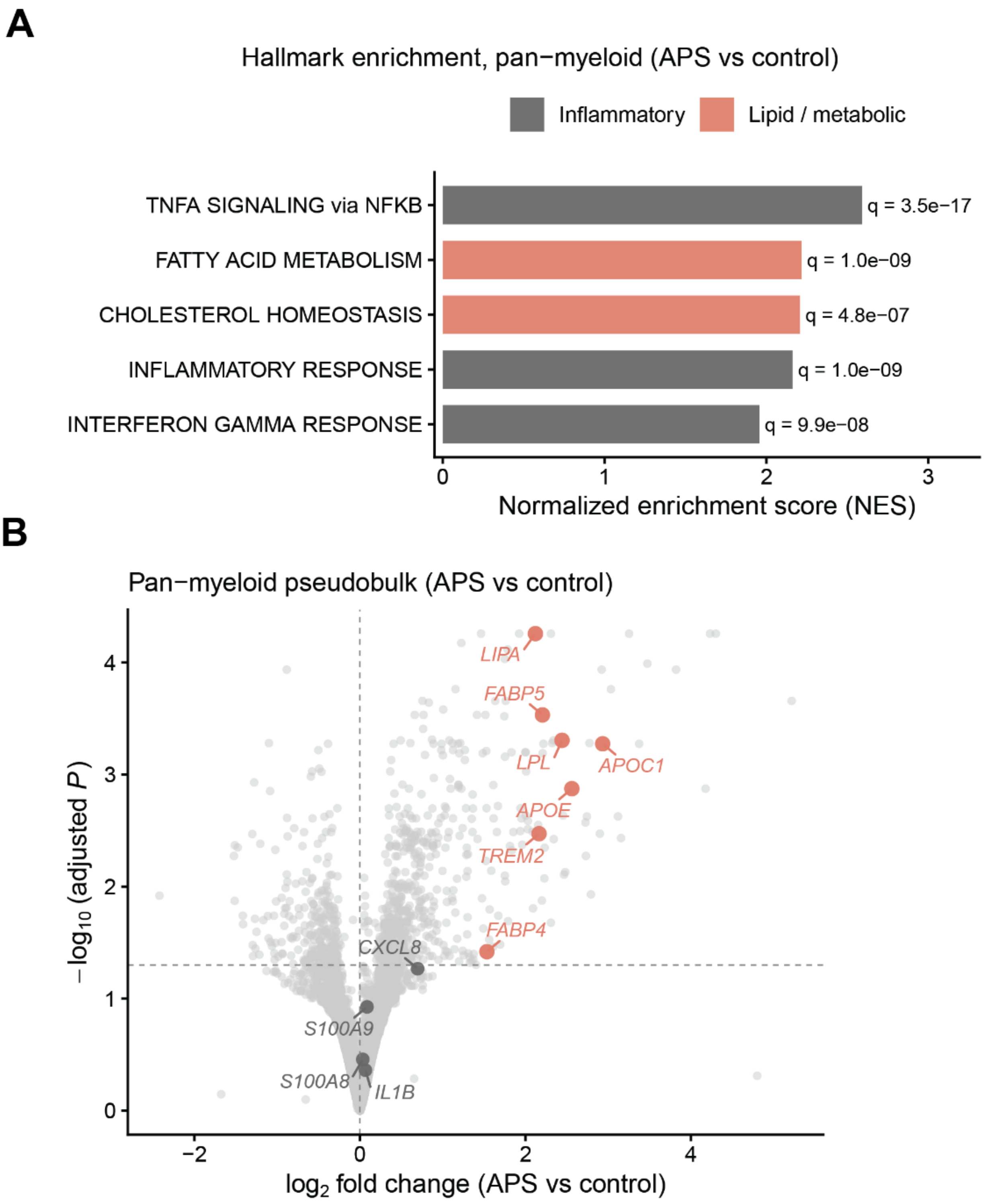


**Figure S1. Pseudobulk transcriptional changes in the APS dermal myeloid compartment.**

(A) Gene set enrichment analysis (fgsea) of the upregulated genes in APS myeloid. Enriched programs include lipid-metabolic (fatty acid metabolism, cholesterol homeostasis) and inflammatory (TNF/NF-κB, interferon gamma) gene sets. Bars indicate normalized enrichment score.

(B) Volcano plot of pseudobulk differential expression between APS and HC across the aggregated dermal myeloid compartment (DESeq2). Genes with BH-adjusted P < 0.05 are highlighted; lipid-handling genes (including APOC1, APOE, LPL, FABP5, FABP4, LIPA, TREM2) are labeled. [30 APS / 14 HC pseudobulk samples; log2FC and padj thresholds.

A

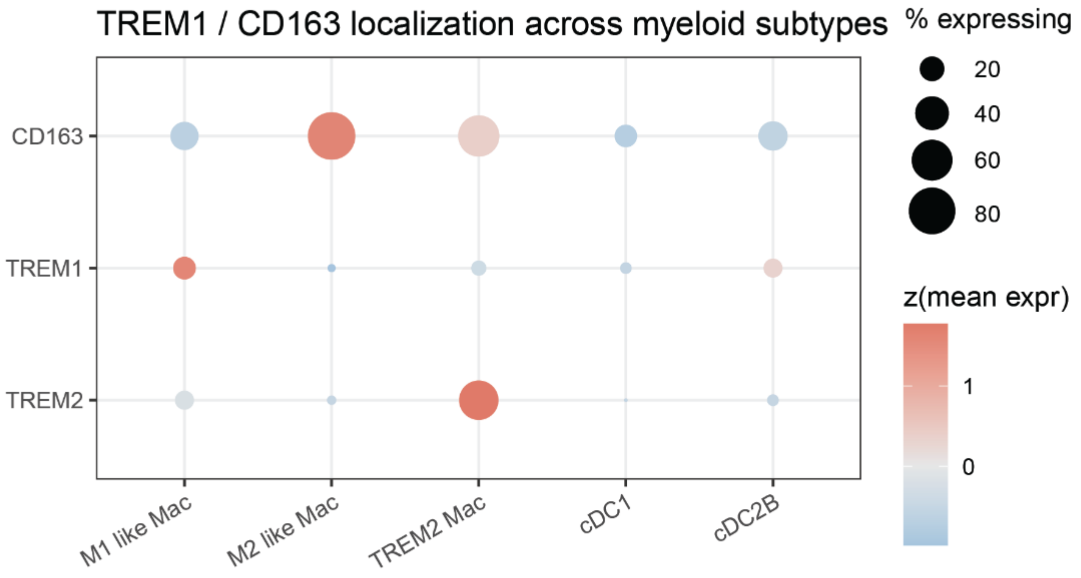


**Figure S2. Molecular identity of the TREM2 macrophage state.**

(A) Dot plot expression of TREM2, the scavenger receptor CD163, and TREM1 across the five dermal myeloid populations, distinguishing TREM2 macrophages ($TREM2^{+}CD163^{+}TREM1^{-}$) from the proinflammatory macrophage phenotype.

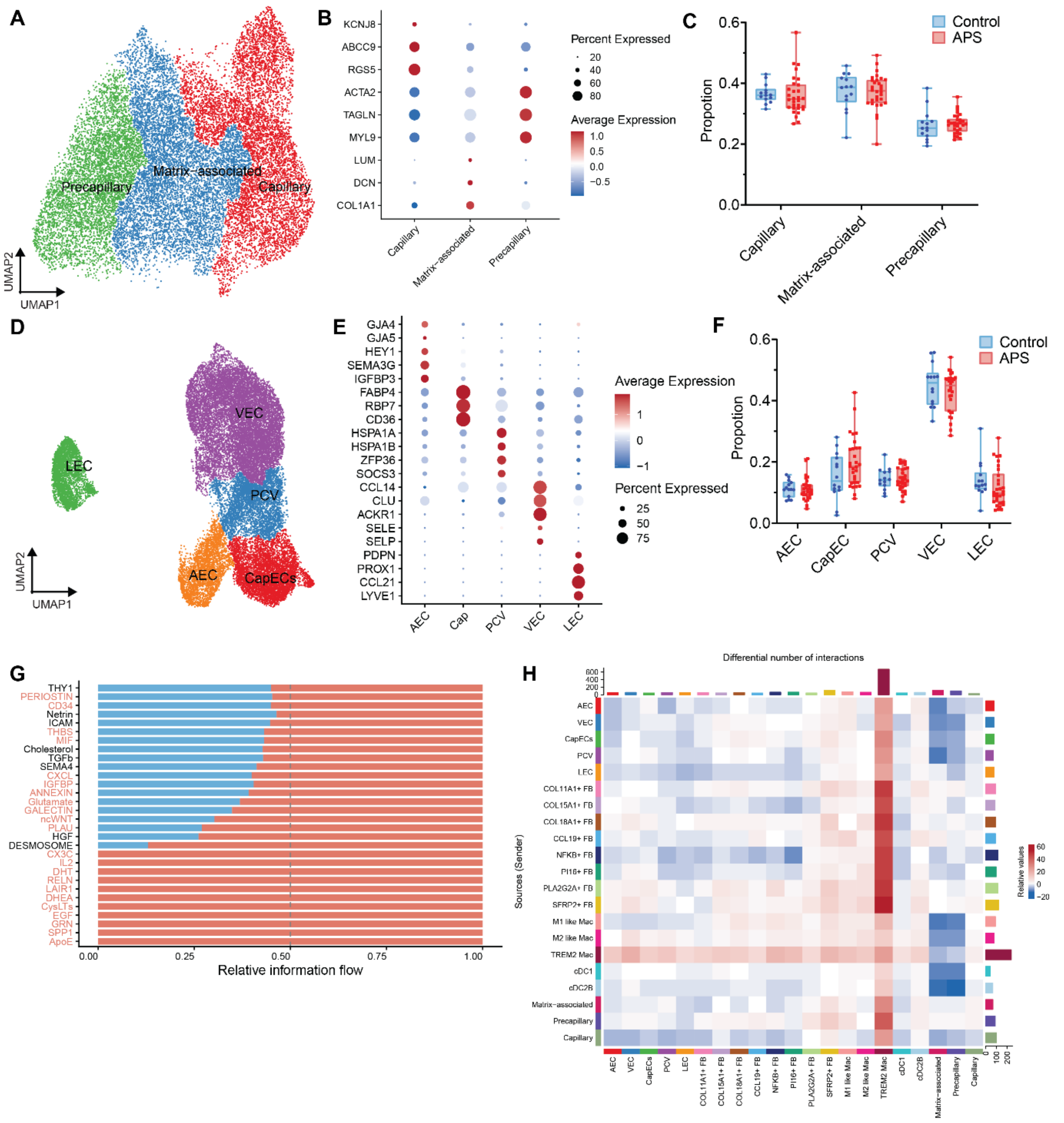


**Figure S3. Endothelial and pericyte subpopulations and the dermal differential-interaction landscape.**

(A) UMAP of the dermal pericyte compartment, resolving matrix-associated, precapillary, and capillary pericyte subpopulations.

(B) Dot plot of representative canonical markers for the pericyte subpopulations.

(C) Sample-level proportions of the pericyte subpopulations in APS and HC, each point is one participant; two-sided Mann–Whitney with BH correction. No subpopulation differed after correction (q > 0.05).

(D) UMAP of the dermal endothelial compartment, resolving five subpopulations: arterial endothelial cells (AEC), capillary endothelial cells (CapECs), post-capillary venule endothelial cells (PCV), venous endothelial cells (VEC), and lymphatic endothelial cells (LEC);

(E) Dot plot of representative canonical markers for the endothelial subpopulations.

(F) Sample-level proportions of endothelial subpopulations in APS and HC, analyzed as in (C). No subpopulation differed after correction ($q > 0.05$).

(G) Ranked information flow of signaling pathways between conditions (CellChat rankNet, stacked). Pathway labels significantly higher in APS by Wilcoxon are indicated in red.

(H) Heatmap showing differential number of predicted intercellular interactions (APS versus HC) across all endothelial, pericyte, myeloid, and fibroblast subpopulations (CellChat compareInteractions, count).

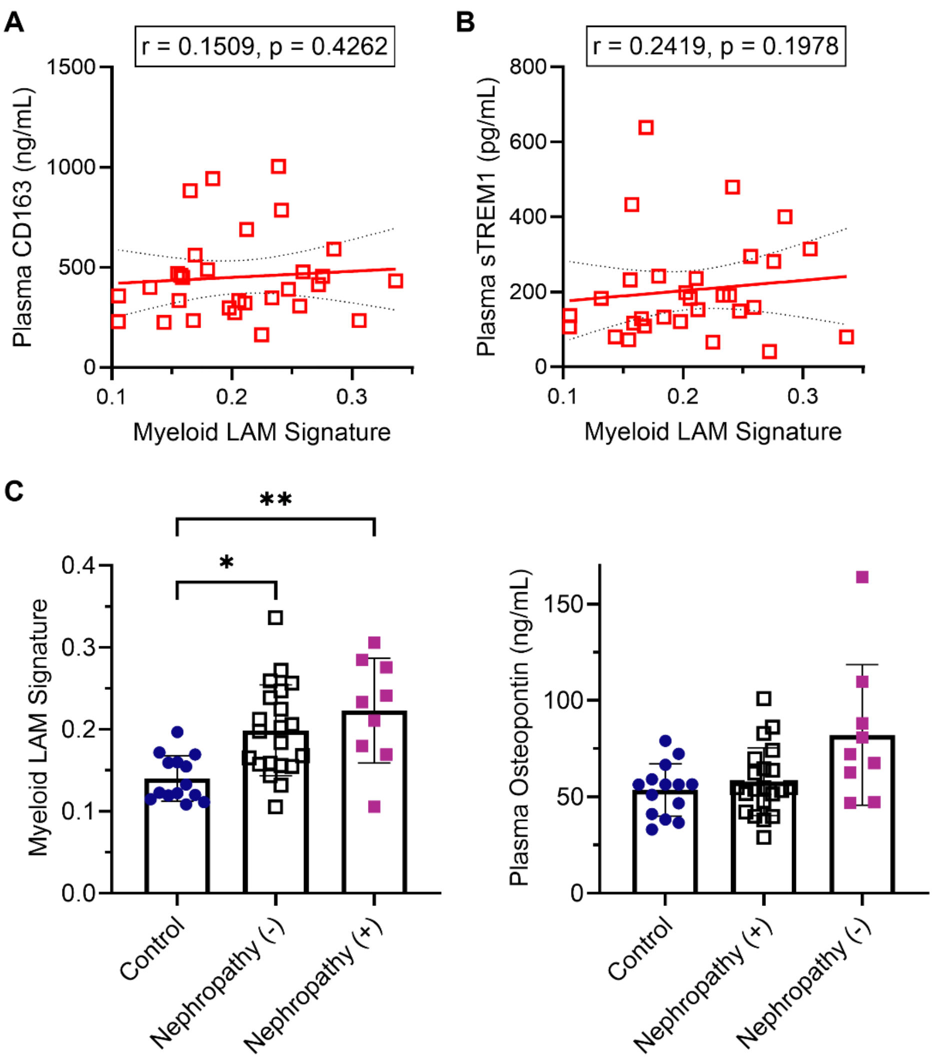


**Figure S4. Plasma sTREM2 is a within-disease correlate independent of macrophage burden, and plasma OPN marks microvascular involvement.**

(A) Association between plasma CD163 and tissue LAM signature among 30 patients with APS, evaluated by two-sided Spearman correlation. Plasma CD163 was not associated with tissue LAM signature (ρ = 0.15; P = 0.43).

(A) Association between plasma sTREM1 and tissue LAM signature among 30 patients with APS, evaluated by two-sided Spearman correlation. plasma sTREM1 was not associated with tissue LAM signature (ρ = 0.24; P = 0.20).

(C) Tissue LAM signature (left) and plasma osteopontin (right) in patients with APS stratified by documented nephropathy (nephropathy+, n = 9; nephropathy−, n = 21). Groups were compared by Kruskal–Wallis test with Dunn's post hoc correction. Neither the tissue LAM signature nor plasma OPN distinguished nephropathy+ from nephropathy− patients.

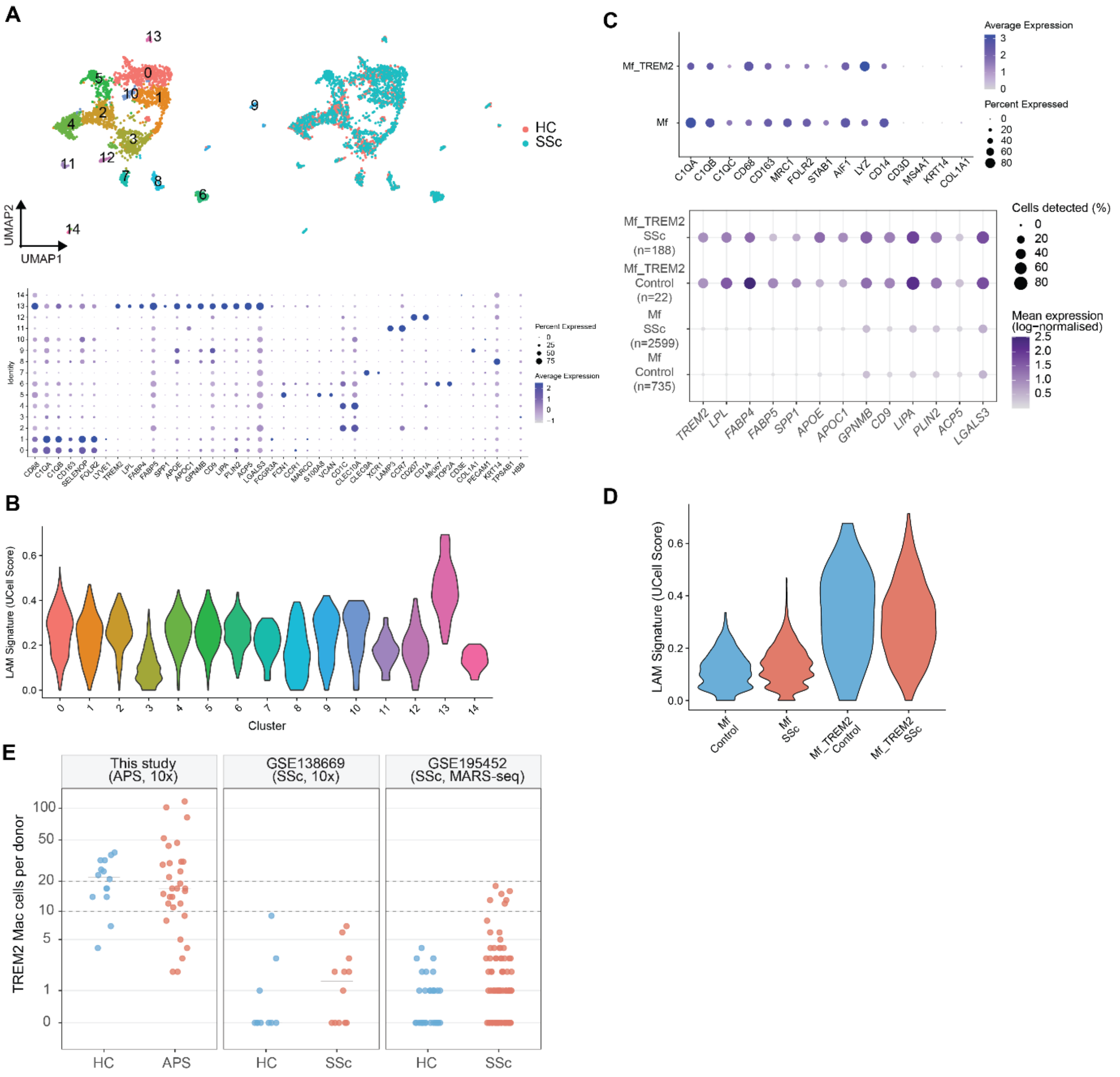


**Figure S5. Reanalysis of two public systemic sclerosis skin single-cell datasets for the TREM2 macrophage program.**

(A) UMAP of the myeloid compartment of GSE138669 (12 dcSSc and 10 HC, forearm skin, 10x), colored by subcluster and by condition, with a dot plot of lineage and core LAM markers across subclusters. Subcluster 13 was annotated as TREM2 Mac based on expression of TREM2, LPL, FABP4, SPP1, APOE, LIPA, PLIN2, and ACP5 with macrophage identity (CD68).

(B) Cell-level LAM-18 signature score (UCell) by myeloid subcluster in GSE138669. Subcluster 13 was the highest scoring population.

(C) Dot plot of macrophage identity markers for the author-annotated Mf and Mf_TREM2 populations of GSE195452 (97 SSc and 56 HC, forearm skin, MARS-seq), and detection of the 13 core LAM genes in the same populations split by disease. Dot size is the percentage of cells with at least one unique molecular identifier (UMI) and color is mean log-normalized expression, no scaled across genes.

(D) Cell-level LAM-18 signature score by population and condition in GSE195452.

(E) TREM2 macrophage cells recovered per donor in this study and in the two SSc datasets, each point is one donor, log(x+1) scale, bars are group medians, dashed lines mark the 10-cell and 20-cell minima used for pseudobulk and effect-size analyses. This study recovered 1,114 cells across 44 donors (median 17, range 2–116), GSE138669 36 cells across 22 donors (median 0, range 0–9), and GSE195452 217 cells across 111 donors (median 1, range 0–18).
